# Transparency and reporting characteristics of COVID-19 randomized controlled trials

**DOI:** 10.1101/2022.02.03.22270357

**Authors:** Philipp Kapp, Laura Esmail, Lina Ghosn, Philippe Ravaud, Isabelle Boutron

## Abstract

Randomized controlled trials (RCTs) are essential to support clinical decision making. We assessed the transparency, completeness and consistency of reporting of 244 reports (120 peer-reviewed journal publications; 124 preprints) of RCTs assessing pharmacological interventions for the treatment of COVID-19 published the first 17 months of the pandemic (up to May 31, 2021). Transparency was poor. Only 55% of trials were prospectively registered; 39% made their full protocols available and 29% provided access to their statistical analysis plan. Only 6% completely reported the most important information. Primary outcome(s) reported in trial registries and published reports were inconsistent in 47% of trials. Of the 124 RCTs published as preprint, 76 were secondarily published in a peer-reviewed journal. There was no major improvement after the peer-review process.

Lack of transparency, completeness and consistency of reporting is an important barrier to trust, interpretation and synthesis in COVID-19 clinical trials.

## BACKGROUND

Due to the global Covid-19 pandemic, clinical research has accelerated dramatically. To date, more than 2900 randomized controlled trials (RCTs) on pharmacological interventions for COVID-19 have been registered [1, 2]. Traditionally time-consuming attributes of clinical trials such as planning, conduct and reporting, have been considerably shortened.

The communication of scientific results has been particularly modified and accelerated to respond to the need and request for rapid information on the COVID-19 from policymakers, guideline developers, health care providers and the public.

Due to the long duration of journal editorial processes, some preprint servers gained up to 25% more trials due to COVID-19 [3]. Medical Journals reacted on the other hand by accelerating their editorial processes by up to 49% on average [4]. Naturally, concerns regarding the quality and transparency of the study results disseminated have been raised [5-10].

We aimed to assess: 1) the transparency, completeness and consistency of reporting in reports of RCTs assessing pharmacologic treatments for COVID-19; and, 2) the impact of the peer-review process on reporting and transparency for all preprints secondarily published in peer-reviewed journals.

## RESULTS

### RCTs identification and characteristics

The results of the search are detailed in Figure 1. Of the 47,061 records screened, 244 reports of randomized trials evaluating pharmacological interventions for the treatment of COVID-19 were identified and assessed. Overall, 120 (49%) RCTs were first published in peer reviewed journals while 124 (51%) were initially available as preprints. Of the 124 preprints, 76 (61%) were subsequently published in a peer-review journal. Table 1 provides information on the general characteristics of the trials. Overall, 89% (n = 218) were conducted in a single country (countries with the most conducted trials were Iran [n = 38], China [n = 31], USA [n = 20] and Brazil [n = 20]). Most RCTs used a 2-arm design (n = 210, 86%). The median sample size was 100 (IQR: 54-245) (range 10 to 11558). Overall, about half of the trials were open-label.

**Table 1.** Randomized trials characteristics.

| Characteristic | Overall<br>n = 244 | Preprints<br>n = 124 | Journal publications<br>n = 120 |
| --- | --- | --- | --- |
| **Number of arms** |  |  |  |
| • 2 arms | 210 (86%) | 106 (85%) | 104 (87%) |
| • More than 2 arms | 34 (14%) | 18 (15%) | 16 (13%) |
| **Sample size (median, IQR)** | 100 (54 – 245)* | 95 (51 – 248)* | 107 (60 – 240) |
| **Setting** |  |  |  |
| • Single country | 218 (89%) | 108 (87%) | 110 (92%) |
| • Multinational | 26 (11%) | 16 (13%) | 10 (8%) |
| **Number of centres** |  |  |  |
| • Single-centre | 101 (41%) | 52 (42%) | 49 (41%) |
| • Multicenter | 139 (57%) | 72 (58%) | 67 (56%) |
| • No information | 4 (2%) | 0 (0%) | 4 (3%) |
| **Blinding** |  |  |  |
| • Any blinding | 107 (44%) | 58 (47%) | 49 (41%) |
| • No blinding | 128 (52%) | 62 (50%) | 66 (55%) |
| • Unclear | 9 (4%) | 4 (3%) | 5 (4%) |

\* Eight trials did not report the sample size (missing data)

### Transparency indicators

#### Access to the trial documentation

A trial protocol was available in 39% (n = 95) of trials, 4 (2%) were not in English. A statistical analysis plan was accessible in only 29% of the trials (n = 71).

#### Trial registration

Overall, 231 trials (95%) were registered; 88 (36%) were retrospectively registered of which 37/88 (42%) had a delay of more than 30 days between the trial start date and registration date. For nine trials (4%) we could not determine if the trial was prospectively posted, due to unclear or missing information in the registry or report.

When the option to post trial results was available (n = 158, 65%), only 27 (17%) posted their results in clinical trial registries (status last checked October 7, 2021). In addition, one trial provided result summary in the registry although posting was not required.

#### Data sharing statement

Overall, 165 trials (68%) made a data sharing statement available in the report. Of those, 150/165 trials (91%) stated their willingness to share data, 14/165 trials (9%) stated that they were not willing to share their data, while one trial (1%) reported to be undecided. Of the 150 trial reports that intended to share data, authors reported that they would share data upon email request (n = 103, 69%) or on an online repository (n = 32, 21%), while 15 trials (10%) did not report how data would be made available.

When a data sharing statement was reported in the registry (n = 170), we identified discrepancies between the data sharing statement in the registry and trial report for 55 trials: from data sharing willingness ‘No’ or ‘Undecided’ in registry to ‘Yes’ in the report (n = 51) and from ‘Yes’ in the registry to ‘No’ in the report (n = 4).

### Completeness of reporting

Results are detailed in Table 2 and S2 Table. Overall, reporting was poor. Only 78 (32%) of the reports completely reported the pre-specified primary outcome; 201 (82%) described the methods used to generate the random allocation sequence and 139 (57%) the process of allocation concealment. Of the studies that reported to be blinded (n = 107, 44%), less than half (n = 43, 40%) clearly described who was blinded and how. About half of the trials (n = 129, 53%) provided a complete description of the participant flow, either as diagram or in text form. Regarding the study results, 124 trials (51%) reported the primary outcome(s) completely. Harm was adequately described in only 15% of the trials (n = 36). In particular, information regarding the mode of harm data collection (i.e., how data was collected) (n = 134, 55%) and the time frame of observation (n = 126, 52%) was insufficiently reported (S2 Table). Fifty-four trials (22%) did not report any results on harms and most trials (n = 147, 60%) did not highlight whether harms resulted in withdrawals or trial discontinuations (S2 Table). Overall, only 6% (n = 15) completely reported the 10 most important CONSORT items.

**Table 2.** Completeness of reporting of CONSORT items and additional variables.

| Item | Checklist item | Complete reporting overall (n = 244) | Complete reporting in preprints (n = 124) | Complete reporting in journal publications (n = 120) | p-value (Fisher's exact test) |
| --- | --- | --- | --- | --- | --- |
| **Consort items** |  |  |  |  |  |
| Outcome | Completely defined pre-specified primary outcome measures, including how and when they were assessed | 78 (32%) | 35 (28%) | 43 (36%) | 0.22 |
| Sequence generation | Method used to generate the random allocation sequence | 201 (82%) | 103 (83%) | 98 (82%) | 0.87 |
| Allocation concealment | Mechanism used to implement the random allocation sequence (e.g., sequentially numbered containers), describing any steps taken to conceal the sequence until interventions were assigned | 139 (57%) | 65 (52%) | 74 (62%) | 0.16 |
| Blinding | If done, who was blinded after assignment to interventions (e.g., participants, care providers, those assessing outcomes) and how | 43/107 (40%) | 21/58 (36%) | 22/49 (45%) | 0.87 |
| Participant flow | For each group, the numbers of participants who were randomly assigned, received intended treatment, and were analyzed for the primary outcome (including losses and exclusions with reasons) | 129 (53%) | 65 (52%) | 64 (53%) | 0.90 |
| Outcomes and estimation | For each primary outcome, results for each group, and the estimated effect size and its precision (such as 95% confidence interval) | 124 (51%) | 63 (51%) | 61(51%) | 1.00 |
| Harms | All-important harms or unintended effects in each group | 36 (15%) | 9 (7%) | 27 (23%) | 0.001 |
| Registration* | Registration number | 223 (91%) | 120 (97%) | 103 (86%) | 0.003 |
| Overall | Overall CONSORT assessment | 15 (6%) | 4 (3%) | 11 (9%) | 0.064 |
| **Additional items** |  |  |  |  |  |
| Funding | Funding information | 231 (95%) | 121 (96%) | 110 (92%) | 0.048 |
| Conflict of interest | Statement of conflicting interests | 230 (94%) | 120 (97%) | 110 (92%) | 0.10 |
| Ethical approval | Statement of ethical approval | 240 (98%) | 124 (100 %) | 116 (97%) | 0.057 |

\*Thirteen trials were not registered (12 peer-reviewed journal publications and one preprint) and nine trials did not present the registration number in the report (Five peer-reviewed journal publications and three preprints)

Most trials reported information on funding (n = 231, 95%). Two hundred-thirty (94%) disclosed information on conflicts of interest. All except four trials (98%) reported ethical approval.

For most items, reporting did not differ between reports first published as preprint and reports first published in a peer-reviewed journal article (Table 2). Results were statistically significant only for the reporting of harm which was better in peer-reviewed journal publication (23% vs 7%, p = 0,001); the reporting was better in preprints for the trial registration number (97% vs 86%, p = 0,003) and funding source (96% vs 92%, p = 0,048)

### Reporting consistency (i.e., primary outcome(s) switching)

Among all registered trials (n = 231), 206 trials (89%) identified their primary outcomes in the report and registry. Of those, 108 (52%) reported primary outcomes as pre-defined in the trial registry. Primary outcome(s) switching between registered and published primary outcome(s) was identified in 97 trials (47%) (Table 3). Switches comprised completely changed primary outcome(s) between report and registry (n = 35, 36%); reports that removed one or several primary outcome(s) (n = 18, 19%); reports that added one or several primary outcome(s) (n = 8, 8%) and reports that added and removed one or several primary outcome(s) (n=16, 17%). In addition, twenty trials (21%) changed the time frame or metric, while the primary outcome variable stayed the same. Twelve trials comprised both, changes in time frames or metrics, as well as added, removed or changed primary outcome(s)

**Table 3.** Primary outcome(s) switching between the registry and report (n = 97)

| **Outcome Switch** | **Example** | **N (%)** |
| --- | --- | --- |
| **Added primary outcome(s)** | Registry:<br>1. Time to clinical improvement<br>Report:<br>1. Death<br>2. Time to clinical improvement | 8 (8%) |
| **Removed primary outcome(s)** | Registry:<br>1. Time and rate of temperature return to normal<br>2. Time and rate of improvement of respiratory symptoms and signs<br>3. Time and rate of change to negative COVID-19 nucleic acid test<br>4. Rate of mild/moderate type to severe type, rate of severe type to critical type<br>Report:<br>1. Rate of nucleic acid negativity conversion of SARS-CoV-2<br>2. Negativity conversion time | 18 (19%) |
| **Added and removed primary outcome(s)** | Registry:<br>1. Hospitalization days<br>2. Need for mechanical ventilation<br>3. Condition of discharge (death or recovery)<br>Report:<br>1. Improvement in the rate of ICU admissions<br>2. Intubation/mechanical ventilation<br>3. Mortality 28-days | 16 (17%) |
| **Changed primary outcome(s)** | Registry:<br>1. Time to improvement<br>Report:<br>2. Clinical status | 35 (36%) |
| **Time frame or metric different** | Registry:<br>1. Change in clinical status of subjects at Day 7<br>Report:<br>2. Change in clinical status of subjects at Day 15 | 20 (21%)* |

Percentages may not add up to 100% due to rounding.
\* Twelve additional trials comprised both, changes in time frames or metrics, as well as added, removed or changed primary outcome(s). Those trials were counted only a single time within the added, removed or changed outcome(s) switching domains.

### Comparison between preprint report and subsequent peer-reviewed journal publication

#### Reports identification

Of 124 preprints included in our analysis, we identified 76 corresponding peer-reviewed journal publications. The median time between preprint and publication in a peer reviewed journal was 95 days (IQR: 59-171) (range: 5 – 505). The protocol and the statistical analysis plan were added in 13 (17%) and 11 (15%) peer-reviewed journal publications respectively. However, the protocol and the statistical analysis plan were also removed respectively in 5 (7%) and 3 (4%) reports.

### Differences in completeness of reporting and primary outcome(s) switching

The detailed differences between preprint and peer-reviewed journal publication are described in Table 4 and S3 Table. Information was rarely added: allocation concealment (n = 6, 8%), the persons who were blinded (n = 5, 7%), the mode of harm data collection (n = 6, 8%) as well as the time frame of harm surveillance (n = 6, 8%). Information that was removed from the preprint in the peer-reviewed journal publication included the mode of harm data collection (n = 4, 5%), the description of the primary outcome (n = 3, 4%) or the registration number (n = 3, 4%). Overall, 40 trials (53%) had at least one change within all CONSORT sub-items. Only two (3%) of the trials had changed their overall CONSORT assessment from partially to completely reported.

**Table 4.** Changes in CONSORT sub-items between preprint and peer-reviewed journal publication (n=76)

| Consort item | Reported no change | Not reported, no change | Added | Subtracted | Not applicable |
| --- | --- | --- | --- | --- | --- |
| **CONSORT Section 6a** |  |  |  |  |  |
| Clear primary outcome | 68 (89%) | 3 (4%) | 2 (3%) | 3 (4%) | 0 (0%) |
| Variable of interest | 67 (88%) | 1 (1%) | 0 (0%) | 0 (0%) | 8 (11%)* |
| How the outcome was assessed | 60 (79%) | 8 (11%) | 0 (0%) | 0 (0%) | 8 (11%)* |
| The analysis metric | 68 (89%) | 0 (0%) | 0 (0%) | 0 (0%) | 8 (11%)* |
| The summary measure for each study group | 56 (74%) | 9 (12%) | 3 (4%) | 0 (0%) | 8 (11%)* |
| Time point of interest for analysis | 61 (80%) | 4 (5%) | 2 (3%) | 1 (1%) | 8 (11%)* |
| Who assessed the outcome | 31 (41%) | 32 (42%) | 4 (5%) | 1 (1%) | 8 (11%)* |
| **CONSORT Section 8a and 9** |  |  |  |  |  |
| Method of sequence generation | 69 (91%) | 4 (5%) | 3 (4%) | 0 (0%) | 0 (0%) |
| Mechanism allocation concealment | 49 (64%) | 20 (26%) | 6 (8%) | 1 (1%) | 0 (0%) |
| **CONSORT Section 11b** |  |  |  |  |  |
| Who was blinded | 24 (32%) | 6 (8%) | 5 (7%) | 0 (0%) | 41 (54%)<sup>†</sup> |
| How the blinding was performed | 30 (39%) | 4 (5%) | 1 (1%) | 0 (0%) | 41 (54%)<sup>†</sup> |
| Similarities of the characteristics of the interventions | 17 (22%) | 11 (14%) | 4 (5%) | 1 (1%) | 43 (57%)<sup>†</sup> |
| **CONSORT Section 13b** |  |  |  |  |  |
| Flow chart | 64 (84%) | 8 (11%) | 4 (5%) | 0 (0%) | 0 (0%) |
| Participants randomized | 73 (96%) | 2 (3%) | 1 (1%) | 0 (0%) | 0 (0%) |
| Participants who received treatment | 67 (88%) | 8 (11%) | 1 (1%) | 0 (0%) | 0 (0%) |
| Participants lost to follow-up | 75 (99%) | 1 (1%) | 0 (0%) | 0 (0%) | 0 (0%) |
| Participants who discontinued intervention | 50 (66%) | 22 (29%) | 3 (4%) | 1 (1%) | 0 (0%) |
| Participants analyzed | 74 (97%) | 2 (3%) | 0 (0%) | 0 (0%) | 0 (0%) |

**Table 4. continued**
| Consort item | Reported<br>no change | Not reported,<br>no change | Added | Subtracted | Not<br>applicable |
| --- | --- | --- | --- | --- | --- |
| **CONSORT Section 17a** |  |  |  |  |  |
| Result | 72 (95%) | 2 (3%) | 1 (1%) | 1 (1%) | 0 (0%) |
| Difference in estimated effect | 48 (63%) | 24 (32%) | 3 (4%) | 1 (1%) | 0 (0%) |
| Precision of the estimated effect | 47 (62%) | 1 (1%) | 0 (0%) | 0 (0%) | 28 (37%)<sup>‡</sup> |
| **CONSORT Section 19** |  |  |  |  |  |
| List of harms addressed | 41 (54%) | 29 (38%) | 4 (5%) | 2 (3%) | 0 (0%) |
| Mode of data collection | 35 (46%) | 31 (41%) | 6 (8%) | 4 (5%) | 0 (0%) |
| Time frame of surveillance | 33 (43%) | 33 (43%) | 6 (8%) | 4 (5%) | 0 (0%) |
| Person responsible making attribution | 28 (37%) | 39 (51%) | 5 (7%) | 4 (5%) | 0 (0%) |
| Participant withdrawals due to harm | 31 (41%) | 42 (55%) | 1 (1%) | 2 (3%) | 0 (0%) |
| Results of each harm type | 65 (86%) | 8 (11%) | 2 (3%) | 1 (1%) | 0 (0%) |
| **CONSORT Section 23** |  |  |  |  |  |
| Registration number | 73 (96%) | 0 (0%) | 0 (0%) | 3 (4%) | 0 (0%) |

Percentages may not add up to 100% due to rounding.
\* Eight publications did not clearly specify the primary outcome (either in preprint or journal publication). Due to that the full assessment of *CONSORT section 6a was (item 2-7)* was not possible.
† Forty-one trials were unblinded and could not be assessed. Two additional trials did not use placebo control and could not be assessed for the item *similar characteristics of the intervention (CONSORT section 11b, item 3)*.
‡ Twenty-eight trials did not present the *difference in estimated effect measure (CONSORT section 17a, item 2)*. Precision was therefore not assessable for 28 trials (*item 3*)

Of the 76 assessed trials, nine trials could not be compared for differences in primary outcome(s) switching, due to the missing definition of the primary outcome in the pre-print (n = 3), peer-reviewed journal publication (n = 2), or both (n = 4). There was no change in primary outcome(s) switching between preprint and peer-reviewed journal publication. Only one trial added a justification for primary outcome(s) switching in the peer-reviewed journal publication.

## DISCUSSION

Our results raise important concerns about the transparency, completeness and accuracy of the reporting of RCTs assessing pharmacological interventions for the treatment of COVID-19. Despite a considerable delay (median 95 days) between the publication of the results on a preprint platform and the publication in a peer-reviewed journal, the peer-review process had no impact on transparency, completeness and accuracy of reporting.

### Comparison with other studies

Our results are consistent with other studies assessing reporting characteristics of RCTs reports [11-14]. An analysis of more than 20.000 RCTs included in Cochrane reviewed showed important deficiencies in reporting which is a strong barrier for risk of bias assessment and the extraction of outcomes needed to conduct systematic reviews and meta-analyses [11, 14]. Primary outcome switching also raises important concerns, of which our results showed higher prevalence compared to other studies performed in other fields [15, 16]. This difference could be explained by the novelty of the disease and the rapid increase in knowledge over time which may have requested important changes to the protocol. Nevertheless, the lack of transparency related to these changes is concerning.

### Strengths and limitations

To our knowledge, this is the first study extensively assessing reporting characteristics of all COVID-19 RCTs assessing pharmacologic treatment published as preprint or peer-reviewed journal article the first 17 months of the pandemic. Our sample has been included in a large living network meta-analysis and is comprehensive. Furthermore, we assessed various dimensions of transparency, reporting and consistency between reports and registry records. Finally, to our knowledge it is the largest study comparing preprint and subsequent publications. Our study has nevertheless some limitations. First, we focused on randomized controlled trials and cannot extrapolate to other study designs. Nevertheless, RCTs are considered the gold standard for therapeutic evaluation. Second, most trials were assessed by a single researcher, however a random sample was extracted in duplicate and showed good reproducibility.

### Implications

Our results have important implications. There is an urgent need for high quality evidence to guide the management of COVID-19 patients. It is consequently essential to improve reporting and transparency and increase adherence to the CONSORT statement. As part of the COVID-NMA living review, we are already systematically contacting investigators to request the missing data. Further, we plan to inform investigators of their results in terms of reporting and transparency to help them improve the content of their reports.

The publication of results on preprint servers became an essential mean of communication. It was adopted considerably by the research community during this pandemic mainly because it shortened delays between the production of reports and their dissemination to the community. In our sample, half of the trials decided to communicate first through preprint. Overall, it reduced the delay of accessing results by a median of 3 months. Some researchers, decision makers, funders, editors raised concerns related to the risk of disseminating reports that were not peer-reviewed [6-8]. However, our results do not support the hypothesis that peer-reviewed journal publications are of better quality compared to preprints. We found no difference in terms of transparency and reporting between the preprint and the peer-reviewed report [17, 18].

Finally, our results question the role of the peer-review process in improving reporting and transparency. Our results are consistent with other studies comparing completeness of reporting of the submitted report to the published report focusing on RCTs [19]. We need to develop specific interventions and tools to increase the detection and improvement of reporting in publications. Some tools such as the CobPeer tool have been proposed and evaluated [20]. Other interventions targeting preprints could be useful to inform trialists of reporting deficiencies and help them improve their report prior publications.

In conclusion, lack of transparency, completeness and consistency of reporting is an important barrier to trust, interpretation and synthesis in COVID-19 clinical trials. Peer-reviewed publications were not better than preprints in this regard. Furthermore, the peer-review process failed to improve the deficiency in reporting.

Trial authors as well as editors and funders must apply higher standards of methodological rigor and transparency to ensure the generation of the highest level of evidence to inform decision-making and curb the pandemic.

## METHODS

### Study design

This study is part of the COVID-NMA initiative [2, 21, 22]. The two first pillars of this initiative are a living mapping and living evidence synthesis of all randomized controlled trials assessing treatments and preventive interventions for COVID-19. All results are updated weekly and made available open access on a platform (https://covid-nma.com) [2, 21, 22].

The third pillar of this initiative described in this manuscript is a monitoring of the transparency, completeness and consistency of reporting in the trial reports.

### Search Strategy

#### Eligibility criteria

We included all RCTs assessing pharmacologic interventions for the treatment of COVID-19. Trials assessing nonpharmacologic interventions (e.g., prone position, physiotherapy), pharmacological treatment of long-COVID and preventive interventions as well as vaccines were excluded. Early-phase clinical trials, single-arm trials, non-randomized studies and modelling studies of interventions for COVID-19 were excluded. There was no restriction on language.

#### Search methods for identification of studies

The search strategy was developed in the context of the COVID-NMA initiative for the living systematic review of all treatment interventions for COVID-19 [21]. In brief, we searched electronic databases daily to identify all COVID-19 randomized controlled trials. The search strategy is detailed in S1 Table. The last search was conducted on May 31, 2021.

For all trials published as a preprint, we systematically searched weekly for a subsequent publication in peer-reviewed journals using a preprint tracker (last search October 7, 2021) [23]. For all references retrieved by the preprint tracker, we selected only the peer-reviewed publication corresponding exactly to the content of the preprint in terms of methods and results reported.

Two reviewers screened all retrieved titles, abstracts in duplicate independently using Rayyan [24]. Discrepancies were resolved by consensus between the two reviewers. A third reviewer was involved to resolve disagreements when necessary.

### Data extraction

We developed a standardized data extraction form covering general trial characteristics, transparency indicators, completeness and consistency of reporting.

To avoid errors during the extraction and ensure calibration, two reviewers were trained and assessed separately 20 studies each. The reviewers discussed the meaning of each assessment item and reached consensus for the 20 studies. Subsequently, all included trials were extracted by a single reviewer. The inter-rater agreement between the two reviewers was good with 96.6% agreement, with a Kappa Coefficient of 0.87.

#### General characteristics of the trials

We extracted the trial design, number of arms, sample size, setting, number of centers and blinding.

#### Transparency indicators

We considered the following indicators of transparency:

1. <u>Access to the trial documentation:</u> We checked whether we had access to the protocol and statistical analysis plan and if it was available in English.
2. <u>Trial registration:</u> We evaluated whether studies were registered, if registration was done prospectively, (i.e., before the initiation of recruitment) and if trial results were posted on the registry where possible (i.e., Clinicaltrials.gov, EU Clinical Trial Register, ISRCTN registry, DRKS – German Clinical Trials Register, jRCT – Japan Registry of Clinical Trials, ANZCTR - Australian New Zealand Clinical Trial registry)[25].
3. <u>Data sharing statement</u>: We recorded whether, when and how investigators planned to share data based on information in trial registration and the report.

#### Completeness of reporting

We evaluated systematically whether the trial report and protocol, if available, adheres to the Consolidated Standards of Reporting Trials (CONSORT) 2010 statement [26]. We decided to focus on 10 CONSORT items which were deemed most important because they are frequently incompletely reported and are necessary for conducting a systematic review to evaluate the risk of bias and record the outcome data [20]. The completeness of reporting was assessed using the COBPeer tool (Table 2 and S2 Table), which considers the CONSORT-items associated with sub-items eliciting what should be reported for each item extracted as stated in the CONSORT 2010 Explanation and Elaboration Explanation paper [20, 27]. Reviewers had to indicate for each sub-item if the requested information was reported (yes/no). Finally, each item was rated as “completely reported”, if all sub-items were adequately reported, “partially reported”, if at least one sub-item was missing and “not reported”, if all-items were missing. For the assessment of the CONSORT items, we systematically considered the primary outcome of the report. If the primary outcome was not clearly identified, we considered the outcome reported in the objective and if none were reported, we assessed the completeness of reporting of all outcomes reported in the publication and recorded the least adequately reported.

In addition to CONSORT related items, we assessed if authors reported information on funding, conflicts of interest for the primary investigators or trial statistician and ethical approval.

#### Consistency of reporting (i.e., primary outcome(s) switching)

We assessed if the first report publicly available was consistent between what was planned and reported in the registry and what was actually reported in the publication. Particularly, we checked for primary outcome(s) switching between the registration and the report. Primary outcome(s) switching was defined as adding, removing or changing an outcome (i.e., the variable of interest, time frame or metric). Trials that failed to provide any timing information in the report or trial registration, were assessed only for a change in the variable of interest. For the assessment of outcome switching, all available registration platforms were used. If the trial registration was modified after the study start date, we considered the latest registration entry before trial start, if available. We checked whether outcome switching was disclosed in the report. Explanations and justifications were considered as valid, as soon authors indicated the changed primary outcomes in the report (e.g., introduction or discussion section of the report).

#### Comparison between preprint reports and related peer-reviewed journal publication

For preprints secondarily published in a peer-reviewed journal, we compared the reporting of the first preprint report available to peer-reviewed publication. Changes between preprint and peer-reviewed journal publication were classified as “added” information (i.e., information missing in the preprint report but reported in the publication), or “subtracted” information (i.e., information reported in the preprint report but removed in the publication) [19].

### Data analysis

The descriptive analysis consisted of frequencies, percentages, and medians with interquartile range. Fisher’s exact test at p < 0.05 was used to compare the reporting between preprint and peer-reviewed journal publication.

## Data Availability

The datasets used and analysed during the current study are available from the corresponding author on reasonable request.

## Acknowledgements

This study as part of the COVID-NMA initiative which received funding from Université de Paris, Assistance Publique Hôpitaux de Paris (APHP), Inserm, Cochrane France (Ministry of Health), the French Ministry of Higher Education and Research, Agence Nationale de la Recherche (ANR), the World Health Organization (WHO).

