## Supplementary material for "Transparency and reporting characteristics of COVID-19 randomized controlled trials": Figure 1

**Figure 1. Flowchart of included randomized controlled trials (RCTs) (last search date May 31, 2021)**

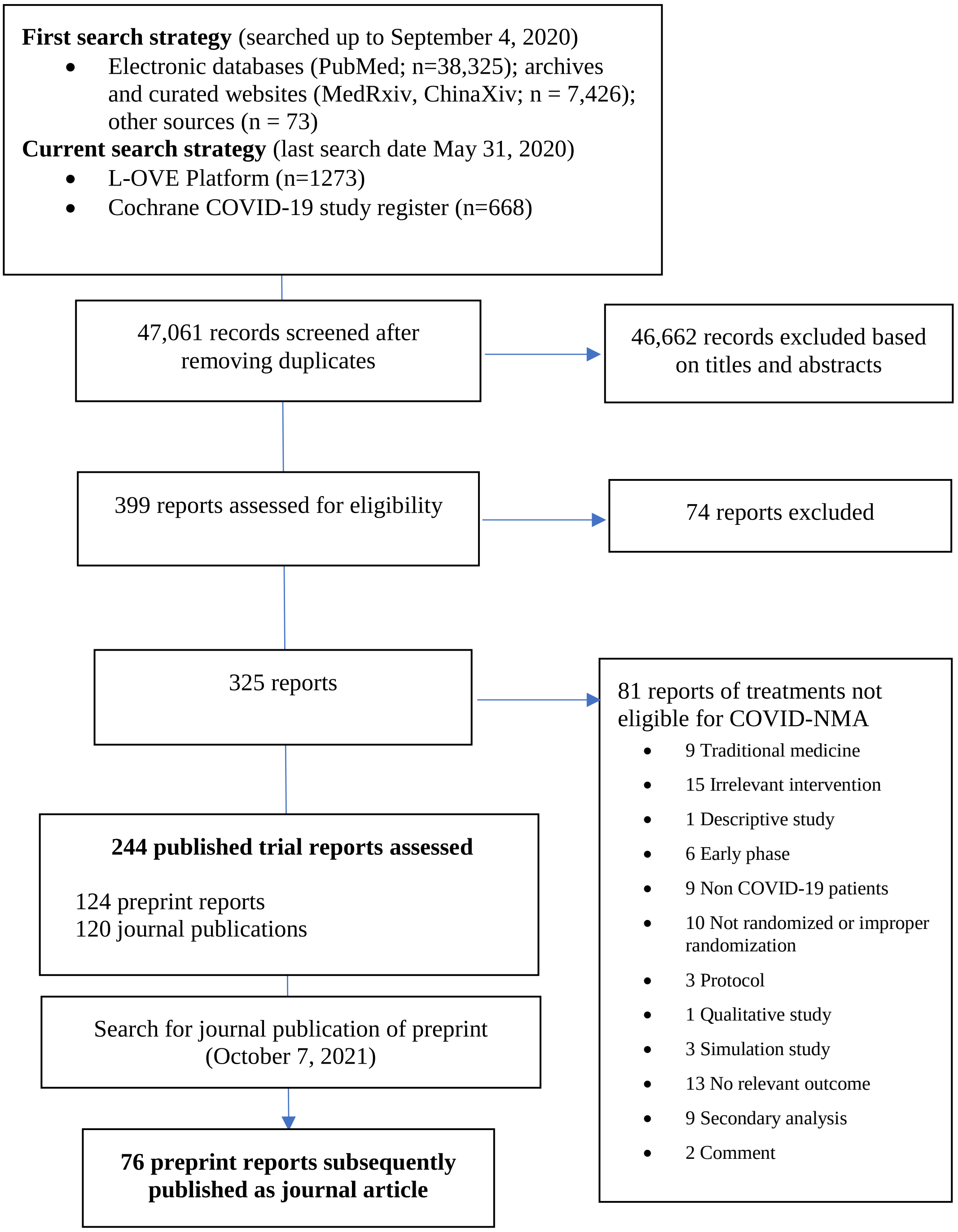


COVID-NMA is a living systematic review of all trials assessing treatment and preventive interventions for COVID-19.
ICTRP: World Health Organization (WHO) International Clinical Trials Registry Platform
