## Supporting Information for "Transparency and reporting characteristics of COVID-19 randomized controlled trials"

**S1 Table. Current search strategy (COVID-NMA)**

| | |
|---|---|
| **PubMed** | (2019 nCoV[tiab] OR 2019nCoV[tiab] OR corona virus[tiab] OR corona viruses[tiab] OR coronavirus[tiab] OR coronaviruses[tiab] OR COVID[tiab] OR COVID19[tiab] OR nCov 2019[tiab] OR SARSCoV2[tiab] OR SARS CoV-2[tiab] OR SARSCoV2[tiab] OR SARSCoV-2[tiab] OR "COVID-19"[Mesh] OR "COVID-19 Testing"[Mesh] OR "COVID-19 Vaccines"[Mesh] OR "Coronavirus"[Mesh:NoExp] OR "SARS-CoV-2"[Mesh] OR "COVID-19"[nm] OR "COVID-19 drug treatment"[nm] OR "COVID-19 diagnostic testing"[nm] OR "COVID-19 serotherapy"[nm] OR "COVID-19 vaccine"[nm] OR "LAMP assay"[nm] OR "severe acute respiratory syndrome coronavirus 2"[nm] OR "spike protein, SARSCoV-2"[nm]) NOT ("animals"[mh] NOT "humans"[mh]) NOT (editorial[pt] OR newspaper article[pt]) |
| **Embase.com** | ((('coronaviridae'/de OR 'coronavirinae'/de OR 'coronaviridae infection'/de OR 'coronavirus disease 2019'/exp OR 'coronavirus infection'/de OR 'SARS-related coronavirus'/de OR 'Severe acute respiratory syndrome coronavirus 2'/exp OR '2019 nCoV':ti,ab,kw OR 2019nCoV:ti,ab,kw OR ((corona* OR corono*) NEAR/1 (virus* OR viral* OR virinae*)):ti,ab,kw OR coronavir*:ti,ab,kw OR coronovir*:ti,ab,kw OR COVID:ti,ab,kw OR COVID19:ti,ab,kw OR HCoV*:ti,ab,kw OR 'nCov 2019':ti,ab,kw OR 'SARS CoV2':ti,ab,kw OR 'SARS CoV 2':ti,ab,kw OR SARSCoV2:ti,ab,kw OR 'SARSCoV 2':ti,ab,kw) NOT (('animal experiment'/de OR 'animal'/exp) NOT ('human'/exp OR 'human experiment'/de))) NOT 'editorial'/it) NOT ([medline]/lim OR [pubmed-not-medline]/lim) AND [1-12-2019]/sd |
| **CENTRAL** | ("2019 nCoV" OR 2019nCoV OR "corona virus*" OR coronavirus* OR COVID OR COVID19 OR "nCov 2019" OR "SARS-CoV2" OR "SARS CoV-2" OR SARSCoV2 OR "SARSCoV-2"):TI,AB AND CENTRAL:TARGET 2 Coronavirus:MH AND CENTRAL:TARGET 3 Coronavirus:EH AND CENTRAL:TARGET 4 #1 OR #2 OR #3 5 2019 TO 2021:YR AND CENTRAL:TARGET 6 #5 AND #4 7 INSEGMENT 8 #6 NOT #7 |

**S2 Table. Completeness of reporting of CONSORT sub-items (N=244)**

| CONSORT Section | Checklist item | Reported N (%) |
|---|---|---|
| **Section 6a** | | |
| Clear primary outcome | Was the primary outcome(s) clearly identified (e.g., the primary/main outcome was pain)? | 219 (90%) |
| Variable of interest | The variable of interest (e.g., pain, all-cause mortality) | 216 (89%) |
| How the outcome was assessed | How the outcome was assessed (e.g., VAS, Beck Depression Inventory score, pain scale) | 189 (77%) |
| The analysis metric | The analysis metric (e.g., change from baseline, final value, time to event) | 214 (88%) |
| The summary measure for each study group | The summary measure for each study group (e.g., mean, proportion with score > 2) | 183 (75%) |
| Time point of interest for analysis | Time point of interest for analysis (e.g., 3 months)**NA if survival analysis* | 195 (80%) |
| Who assessed the outcome | Who assessed the outcome (e.g., the patient, doctor, nurse, caregiver, other) | 94 (39%) |
| **Section 8a and 9** | | |
| Method of sequence generation | Method used to generate the random allocation sequence | 201 (82%) |
| Mechanism allocation concealment | Mechanism used to implement the random allocation sequence (e.g., sequentially numbered containers), describing any steps taken to conceal the sequence until interventions were assigned | 139 (57%) |
| **Section 11b (N=107)** | | |
| Who was blinded | Was the study blinded? | 74 (69%) |
| How the blinding was performed | Who (i.e., participants, healthcare providers, data collectors, outcome adjudicators, and data analysts) was blinded to treatment assignments? | 87 (81%) |
| Similarities of the characteristics of the interventions | How was the blinding performed? (e.g., used of placebo, intervention by physician unaware of the study) | 49 (46%) |

**S2 Table. (continued).**

| CONSORT Section | Checklist item | Reported N (%) |
|---|---|---|
| **Section 13b** | | |
| Flow chart | Did the authors report a flow chart | 204 (84%) |
| Participants randomized | Number of participants randomized in each group | 233 (95%) |
| Participants who received treatment | Number of participants who received intended treatment in each group | 204 (84%) |
| Participants lost to follow-up | Number of participants lost to follow-up with reasons in each group | 236 (97%) |
| Participants who discontinued intervention | Number of participants who discontinued intervention with reasons in each group | 150 (61%) |
| Participants analyzed | Number of participants analyzed for the primary outcome in each group | 235 (96%) |
| **Section 17a** | | |
| Result* | Result in each group (mean (SD) or number of events/N) | 223 (91%) |
| Difference in estimated effect | Difference in estimated effect between groups (e.g., odds ratio (OR), risk ratio (RR), risk difference (RD), hazard ratio (HR), difference in median survival time, mean difference (MD)) | 126 (52%) |
| Precision | Precision for difference between groups (e.g., 95% CI) | 125 (51%) |
| **Section 19** | | |
| List of harms addressed | How harms-related information was collected (List of harm related events that were addressed) | 132 (54%) |
| Mode of data collection | Mode of data collection (Full description of methods used to collect the harm related information) | 110 (45%) |
| Time frame of surveillance | Timing (description of time frame of surveillance) | 118 (48%) |
| Person responsible making attribution | Attribution methods (Person responsible making attribution disclosed and whether blinding was used) | 94 (39%) |
| Participant withdrawals due to harm* | For each group, participant withdrawals due to harm | 96 (39%) |
| Results of each harm type* | Results in each group for each harms type with denominator (mean [SD] or number of events/N) | 189 (77%) |
| **Section 23** | | |
| Registration number | The registration number | 223 (91%) |

* One trial not assessed, since only baseline data were presented. None of the pre-specified outcomes presented in the result section.

**S3 Table. Changes in CONSORT items between preprint and peer-reviewed journal publication (n=76)**

| Consort section | Completely reported, not changed | Partially reported, not changed | Not reported, not changed | Changed to completely reported from partial | Changed to completely reported from not reported | Changed to partially reported from not reported | Changed to partially reported from complete | Changed to not reported from complete | Changed to not reported from partial |
|---|---|---|---|---|---|---|---|---|---|
| **Outcome** | 26 (34%) | 38 (50%) | 3 (4%) | 4 (5%) | 0 (0%) | 2 (3%) | 0 (0%) | 1 (1%) | 2 (3%) |
| **Sequence generation** | 69 (91%) | - | 4 (5%) | - | 3 (4%) | - | - | 0 (0%) | - |
| **Allocation concealment** | 49 (64%) | - | 20 (26%) | - | 6 (8%) | - | - | 1 (1%) | - |
| **Blinding*** | 15 (20%) | 12 (16%) | 1 (1%) | 6 (8%) | 0 (0%) | 0 (0%) | 1 (1%) | 0 (0%) | 0 (0%) |
| **Participant flow** | 43 (57%) | 27 (36%) | 0 (0%) | 5 (7%) | 0 (0%) | 0 (0%) | 1 (1%) | 0 (0%) | 0 (0%) |
| **Outcomes and estimation** | 47 (62%) | 22 (29%) | 2 (3%) | 3 (4%) | 0 (0%) | 1 (1%) | 1 (1%) | 0 (0%) | 0 (0%) |
| **Harms** | 5 (7%) | 59 (78%) | 2 (3%) | 6 (8%) | 0 (0%) | 0 (0%) | 1 (1%) | 0 (0%) | 3 (4%) |
| **Registration** | 73 (96%) | - | 0 (0%) | - | 3 (4%) | - | - | 0 (0%) | - |
| **Overall** | 4 (5%) | 70 (92%) | 0 (0%) | 2 (3%) | 0 (0%) | 0 (0%) | 0 (0%) | 0 (0%) | 0 (0%) |

Percentages may not add up to 100% due to rounding.
* 41 trials were unblinded
